# Estimation and Removal of spurious echo Artifacts in single-voxel MRS using Sensitivity Encoding

**DOI:** 10.1101/2020.09.09.20191460

**Authors:** Adam Berrington, Michal Považan, Peter B Barker

## Abstract

**Purpose:** In localized MR spectroscopy, spurious echo artifacts commonly occur when unsuppressed signal outside the volume-of-interest is exicted and refocused. In the spectral domain, these signals often overlap with metabolite resonances and hinder accurate quantification. Since the artifacts originate from regions separate from the target MRS voxel, this work proposes that sensitivity encoding based on receive coil sensitivity profiles may be used to separate these signal contributions.

**Methods:** Numerical simulations were performed to explore the effect of sensitivity encoded separation for unknown artifact regions. An imaging-based approach was developed to identify regions that may contribute to spurious echo artifacts, and tested for sensitivity-based unfolding of signal contribution on 6 datasets from 3 brain regions. Spectral data reconstructed using the proposed method (‘ERASE’) were compared to standard coil combination.

**Results:** The method was able to fully separate metabolite and artifact signals if regions were known *a priori*. Mismatch between estimated and actual artifact locations reduced the efficiency of artifact removal. Water suppression imaging (WSI) was able to identify unsuppressed signal remote from the MRS voxel in all cases, and ERASE reconstruction (of up to 8 distinct locations) led to improvements in spectral quality and reduced fitting errors for the major metabolites compared to standard reconstruction, without significant degradation of spectral SNR.

**Conclusion:** The ERASE reconstruction tool was demonstrated to reduce spurious echo artifacts in single voxel MRS. ERASE may be incorporated into standard MRS workflows to improve spectral quality when scanner hardware limitations or other factors result in out-of-voxel signal contamination.

## Introduction

Robust quantification of metabolite concentrations in single-voxel (SV) MR spectroscopy (MRS) of the brain is critically dependent on spectral quality. However, spectra are often observed to contain a number of artifacts (1) which can lead to poor metabolite fitting,rejection of datasets based in quality criteria, and ultimately lowers the value of MRS for either research or clinical use. One commonly observed artifact is the spurious echo. In the spectral domain, the spurious echo manifests as an oscillatory or ‘beating signal’ spanning a range of frequencies and often overlaps with metabolite resonances and is also referred to as ‘ghosting artifact’. A rigorous description of the origin of spurious echoes in MRS has been provided in terms of coherence pathways (2) and later k-space formalism within a PRESS localization sequence (3). Essentially, spurious echoes arise from spins outside the voxel of interest which are excited at some point during the pulse sequence, e.g. due to imperfections in slice localization and coherence pathway selection, and then subsequently refocused by sequence elements or magnetic field gradients, generating an echo during signal reception. On Fourier transformation, this signal usually has a large first order phase error because it occurs at some variable time point in the free induction decay. Given the sizeable concentration difference between water and metabolites in ^1^H-MRS of brain (∼10^4^), the main contribution to the spurious echo artifact arises from regions of unsuppressed water signal (3), although lipid resonances may also form spurious echoes.

It has been shown that when slice localization overlaps with air-filled cavities such as the frontal sinuses, the likelihood of observing spurious echo artifacts is increased (4). Susceptibility differences at these air-water interfaces causes local B_0_-field perturbation and hence the water resonance can be shifted outside of the bandwidth of the water suppression pulses. It has also been shown that localized higher-order shimming on small volumes-of-interest, as is commonly performed in MRS, generates an inhomogeneous B_0_ field outside the VOI which leads to regions of poor water suppression (4).

A number of methods to remove these artifacts have been proposed. The use of outer-volume suppression pulses (5) can reduce the artifact intensity, as well as improved RF pulses for spatial localization (i.e. minimizing excitation of out-of-voxel magnetization) (6,7). Phase cycling schemes are used to eliminate unwanted coherence pathways and therefore reduce presence of these artifacts. However, phase cycling is often performed with a small number of steps and is susceptible to subject motion. Crusher gradients also dephase unwanted coherences, however their efficacy may be limited if gradient strengths (and/or durations) are constrained by scanner hardware or sequence design (8). An early study showed that the order of the slice selective pulses (and hence cumulative crusher effect) could be used to minimize artifact intensity (4), although the optimum order would likely vary depending on the voxel location and subject. Recent work has introduced mathematically optimized crusher schemes for different localization sequences (DOTCOPS) (9). In this way, unwanted coherences could be reduced. Nevertheless, crushing power is always limited by hardware constraints, acquisition geometry and the minimum TE desired. Recently, a method to remove spurious echoes in MR spectra has examined the use of deep learning techniques, which were trained on simulated datasets (10). The flexibility of applying deep learning to *in vivo* datasets, however, depends on the nature and extent of the training data.

The separation of aliased signal components based on their spatial position has been demonstrated in MRS using sensitivity-based techniques akin to SENSE acceleration (11,12). One study showed that lipids aliased into a spectroscopic imaging grid could be separated from brain metabolite signals using coil sensitivity information (13). In another study, after performing dual-voxel excitation, reconstruction of separate signals from the left and right hemispheres could be performed based on the sensitivity weighting of the receive coils, thereby accelerating the MRS acquisition by a factor of 2 compared to the sequential acquisition of spectra from each hemisphere (14). It has also been previously proposed to add phase-encoding gradients to SV MRS acquisitions followed by spatial Fourier transformation, in order to separate the desired ROI signal from out-of-voxel artifacts (15–17).

In the current study, it was hypothesized that sensitivity information from multiple receive coils could be used to separate spurious echo artifacts from metabolite signal during a single-voxel acquisition, given that spurious echoes arise from regions outside of the volume-of-interest. A requirement for this method is that the location of the artifact region must be known in order to successfully separate it from the actual region of interest. Here, potential regions of artifact (regions with unsuppressed water signal) were estimated by prepending a water suppression module to a conventional imaging sequence. The combination of water suppression imaging with SENSE MRS reconstruction is termed Estimation and Removal of Artifacts using Sensitivity Encoding (‘ERASE’).

## Theory

To separate spurious echoes arising from distinct spatial locations from the desired signal, the SENSE unfolding technique is used (11,13,14). In a *K*-channel receive array, the time-domain signal measured in the *k^th^* channel, *X_k_*, can be expressed as a weighted sum of the desired signal, *x_v_*, from a voxel, *V*, and *N* artifact signals, *x_j_*, assumed to arise from *N* separate regions, such that

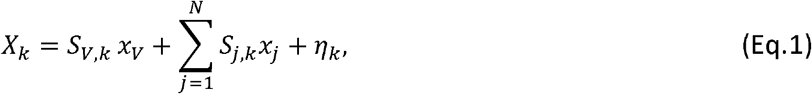

where the weighting factor, *S_i,k_*, is the complex receive sensitivity of the *k^th^* receive channel in region, *i*, and η*_k_*, is the noise observed in channel *k*. By extending (Eq. 1) to a linear system of equations for all *K* receive channels and solving for *x*= (*x_V_*, *x_1_*… *x_N_*), the N+1 aliased signals can be unfolded using the pseudoinverse of the sensitivity matrix, S, such that

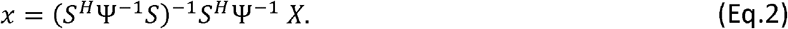

Here, Ψ is the *K x K* noise covariance matrix of the receive array and *S^H^* is the conjugate transpose. Effective separation of multiple signals using receive coil sensitivities relies on a unique weighting of each receive channel, at each location. Regions with similar channel sensitivities lead to poorly-conditioned signal unfolding and noise amplification which is dependent on the coil geometry. The SNR penalty of unfolding relative to separate reconstruction of each region, *i*, is expressed as the geometry factor (g-factor), *g*, such that,

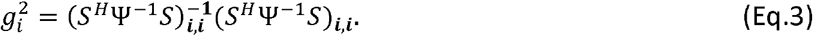

Eq. 3 is identical to previous SENSE approaches to multi-voxel MRS (14) since the unfolding of artifact and metabolite signals can be considered as a form of accelerated parallel reconstruction of simultaneously excited signals.

## Methods

### Simulation of artifact removal

In order to assess the feasibility of the proposed sensitivity-based artifact removal method, simulations of SV MRS acquisition with the addition of spurious echo artifact were performed. A set of complex sensitivity maps were simulated using the Biot-Savart law using a freely available toolbox for parallel MRI written in MATLAB (MathWorks, Natick, MA, USA) (18). Signal reception was simulated using a 16-channel circular array of 30 cm diameter, and 4 cm coil radius, over a 2D spatial grid of 256×256 mm^2^ with 1×1 mm^2^ resolution. A ‘ground truth’ metabolite signal was modelled by density matrix simulations for a STEAM localized acquisition (TE = 14 ms) arising from a 2×2cm^2^ voxel. A single artifactual signal component was simulated as an echo-based complex exponential signal at a chemical shift value of 3.9 ppm and with a refocused echo occurring 164 ms after the start of signal reception. The off-diagonal components of the simulated noise covariance were zero. For the complete list of simulation parameters see Supporting Information S1.

For data acquired *in vivo*, the spatial origin of spurious echo artifacts may not be known accurately, so it is important to understand how mismatch between the estimated and actual artifact location affects the performance of the ERASE method. Therefore, simulations were performed wherby the artifact reconstruction region was systematically varied across the 2D spatial grid, while keeping the position of origin of artifact fixed in addition to the MRS voxel. At each grid position, the percentage of artifact removed (removal (%); defined below) was calculated. A total of 14,017 reconstructions were performed (excluding voxels at the boundary of the image and those overlapping with the ‘known’ MRS location).

The ‘ground-truth’ artifact-free metabolite signal was subtracted from the metabolite signal obtained using ERASE reconstruction at each location to generate the signal, *E*, containing only residual artifact in the reconstructed spectra in addition to noise. The standard multi-channel combined spectrum was calculated and the artifact signal in this spectrum was estimated, *S*. The degree of artifact removal was assessed using a removal score, calculated relative to the standard reconstruction, by comparing the standard deviations of the magnitude of these signals at each grid position in the simulation such that,

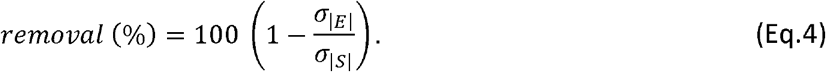

The standard deviation was chosen to account for any difference in DC offsets in the reconstructed signals. Thus, using (Eq. 4) a removal score of 100% indicates, σ_|*E*|_ = 0 i.e.no residual artifact signal in the ERASE reconstructed spectrum and a score of 0% indicates that the level of artifact signal in ERASE spectrum is the same as that in standard reconstruction.

### MRS acquisition

Experimental data was acquired with five healthy volunteers (mean age = 33 years, 1 female, 4 male) on a 3T Philips Ingenia Elition MR system with 32-channel receive array head coil. A total of 6 datasets (R01-06) were obtained from three brain regions; two in occipital cortex (OCC; 15×15×15 mm^3^, R04,R06), one in putamen (30×15×15 mm^3^, R02) and three in anterior cingulate cortex (ACC; 20×20×20 mm^3^ for R03,R05 and 15×15×15 mm^3^ for R01). A T_1_-weighted MPRAGE acquisition (TR = 13 ms, TE = 3.5 ms, 1 mm in-plane resolution) was additionally acquired for voxel placement.

MR spectra were acquired using PRESS localization (TE = 30 ms, TR = 2 s) with VAPOR water suppression (bandwidth = 100 Hz). For test purposes, in order to increase the presence of spurious echo artifacts, no phase cycling was performed. In addition, projection-based shimming was carried out using up to 2^nd^ order corrections, optimized over the same spatial extent as the observed voxel; this is known to increase field inhomogeneity in parts of the sample remote from the voxel, leading to increased spurious echoes. A total of 64 transients were acquired.

For assessment of the efficacy of the proposed ERASE method, comparisons were made to conventionally reconstructed multi-channel spectra (here termed ‘standard reconstruction’), which were calculated using the coil sensitivity information at the target voxel and the measured noise covariance (equivalent to a coil sensitivity-based weighted combination). After coil-combination, individual transients were frequency and phase-corrected before being averaged.

### Calculation of coil sensitivities

Individual channel sensitivity maps were determined using a 3D gradient echo acquisition (TE/TR =0.97/4.1 ms, 3.5 mm isotropic resolution, acquisition time 43 s). Complex channel sensitivities were calculated using ESPIRiT (19) as part of the BART Toolbox for computational MRI (20), using the raw k-space data. The complex noise covariance between the 32 receive channels was calculated using noise samples measured on each channel before acquisition of the gradient echo image data. For the unfolding of spectral and artifact signals from multiple regions (Eq. 2), coil sensitivities were calculated as the arithmetic mean of the sensitivity over the MRS voxel or artifact ROIs.

### Spectral fitting

To compare the performance of artifact removal using either the ERASE method or standard coil combination, spectra were fit using LCModel (21). A basis set was generated using 2D density matrix simulations of PRESS acquisition (TE = 30 ms) of 18 commonly observed metabolites, namely: alanine, ascorbate, aspartate, creatine, GABA, glutamine, glutamate, *myo*-inositol, lactate, phosphocholine, phosphocreatine, phenylethyanolamine, *scyllo*-inositol, taurine, glucose, glycerophosphocholine, glutathione, N-acetylaspartate and N-acetylaspartyl-glutamate. Since estimated concentration values were not the focus of this study, no corrections for T_2_ relaxation times were applied.The Cramér–Rao Lower Bounds (CRLBs) of fitting were compared using both coil combination methods. Subjects R04, R03 and R06 were fit over the range 1.8 to 4.2 ppm due to severe lipid contamination, whereas other subject’s spectra were fit over the range 0.5 to 4.2 ppm.

Raw SNR was measured in the spectral domain as the height of the NAA peak at 2.01 ppm divided by the standard deviation of the noise in the range 15 – 20 ppm using the FID-A toolbox (22). In addition, LCModel reported SNR values were compared between ERASE and standard coil combinations.

### Water-Suppression Imaging (WSI)

The artifact is believed to originate in most cases from regions of the head where B_0_-field inhomogeneity has shifted the water resonance outside of the bandwidth of the water suppression pulses. Thus, in order to identify the location of the artifact signal *in vivo*, water suppression imaging (WSI) maps were acquired using a modified sequence similar to that described in reference (4). WSI maps were acquired after MRS using a 2D turbo spin echo sequence (64 slices, 3.5mm isotropic resolution, TE = 160 ms, TR = 3s, acquisition time 2 m 27 s) and included a VAPOR water suppression preparation module (RF pulse bandwidth = 100 Hz). Importantly, WSI was performed with the identical first- and second-order shim settings, in addition to water suppression parameters, as those used to acquire the MRS signal. In addition, a whole-brain B_0_ fieldmap (TR = 8.2, TE = 3.3 and 5.6 ms, 3.5 mm isotropic, acquisition time 1 m 6 s) was acquired after MRS acquisition using identical shim settings for validation of the WSI method.

### Estimation of artifact regions

In order to determine regions which are solely influenced by water suppression, and to remove tissue contrast from the images, a residual WSI (rWSI) approach was developed by normalizing the WSI images to an identical acquisition acquired without water suppression (non-WSI). An automated algorithm was developed to subsequently partition binary thresholded rWSI maps (thresholded at the mean image intensity of non-zero voxels) into separate artifact regions for reconstruction. Regions were segmented and labelled by iteratively eroding the rWSI maps and then applying a watershed transform (23) to generate unique contiguous regions based on nearest neighbor connectivity. The region segmentation algorithm was implemented in MATLAB (MathWorks, Natick, MA, USA). This iterative process continued until fewer than 8 possible artifact regions were identified. Regions were also constrained to be bigger than a single image voxel however no constraint was placed on the maximum region size.

## Results

### Simulation of artifact removal perfomance

Results from the simulations are shown in Fig. 1. The numerical phantom contained a simulated metabolite signal originating from MRS region (V) and a simulated spurious echo artifact originating 5 cm from the metabolite region (Figure 1A) with a spectral profile centered on 3.9 ppm (Fig. 1A and B). Standard reconstruction, using sensitivity-weighted combination, resulted in considerable contamination of the metabolite spectrum by the spurious echo. Fig. 1C shows the removal performance (Eq. 4) using the ERASE method across the numerical phantom grid, performed on region V and every other point in the grid. Contour lines indicate spatial positions with equal removal performance. Fig. 1D shows the ERASE-reconstructed metabolite and artifact spectra at three different spatial locations in the grid (a – c). Region (a) is centered exactly on the true simulated region (A) and there was complete separation of artifact and metabolite spectra using ERASE, noted by the flat residual signal when compared to the simulated spectrum (removal score = 95%).As the reconstruction region moved further from (A) the removal performance deteriorated. Reconstructing signal at region (b) (distance from A = 8.5 cm), resulted in 60% removal of the simulated artifact from metabolite signal although there was no overlap with the true artifact region. The ERASE reconstructed metabolite spectrum was visibly improved relative to standard reconstruction (Fig. 1B). In extreme cases, when the reconstruction region was positioned on the opposite side of the MRS voxel to the true artifactual region, ERASE performed worse than standard combination shown in region (c) (removal score = −32 %). These simulations illustrate the importance of accurately identifying the location of the artifact signal in order for it to be successfully eliminated using ERASE.

**Fig.1:**
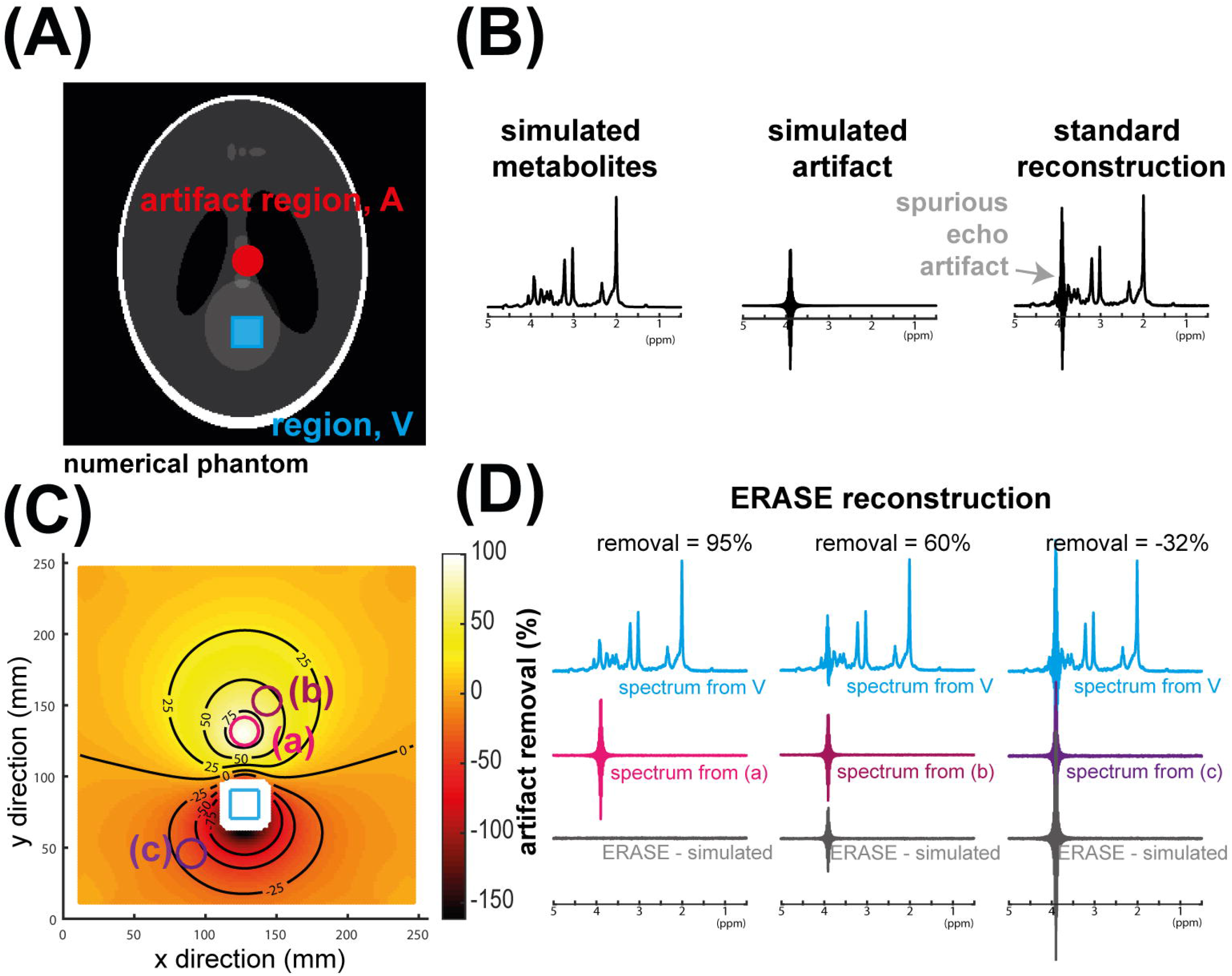
Numerical phantom used to simulate the proposed artifact removal method. **A**: MRS region (blue) and artefact region (red) shown overlaid onto phantom containing 16 complex sensitivity maps [1]. **B**: The simulated metabolite spectrum (STEAM, TE = 14ms) and spurious echo artifact spectrum were placed in the phantom in (A). **C**: Heatmap showing the removal performance using ERASE method when the artifact reconstruction region varied over the whole simulation grid. **D**: Example reconstructed spectra at 3 spatial points in the phantom (a-c).

### Water suppression imaging (WSI) and artifact region estimation

For the estimation of spatial origin of artifacts *in vivo*, water suppression imaging (WSI) maps were acquired in each volunteer. The WSI maps and corresponding residual water suppression imaging (rWSI) maps, showing regions of unsuppressed water, are shown for two volunteers in Fig 2. WSI, acquired using identical second order shims to the MRS acquisition, was found to very clearly show regions of poor water suppression indicated by higher signal intensity. In all acquisition geometries, there was unsuppressed signal in the oral cavity and adjacent structures, infratentorial regions and other regions distal from the MRS acquisition voxel. Peri-cranial lipid signal was visible in all WSI images, since it lies outside the water suppression bandwidth and is unaffected by the water suppression module. Placing the MRS voxel in an anterior region (Fig. 2A) led to large regions of unsuppressed water in the cerebellum and above the frontal sinus. Conversely, for the voxel positioned in the occipital region (Fig. 2B), the frontal lobe contained a large area of unsuppressed water. The spatial distribution of the WSI images was observed to correspond strongly with the B_0_ distribution after localized shimming. When normalized to the image acquired without water suppression and masked, rWSI maps could accurately reveal unsuppressed regions, without the underlying grey and white matter contrast observed in the WSI maps themselves.

The automated thresholding procedure used in this work, resulted in separate contiguous regions of potential spurious echo artifact for each MRS region. The algorithm led to estimation of between 2 and 8 regions. There was a large variation in estimated region volume (mean ± standard deviation: 57±111 cm^3^, minimum: 0.086 cm^3^, maximum: 573 cm^3^).

**Fig. 2:**
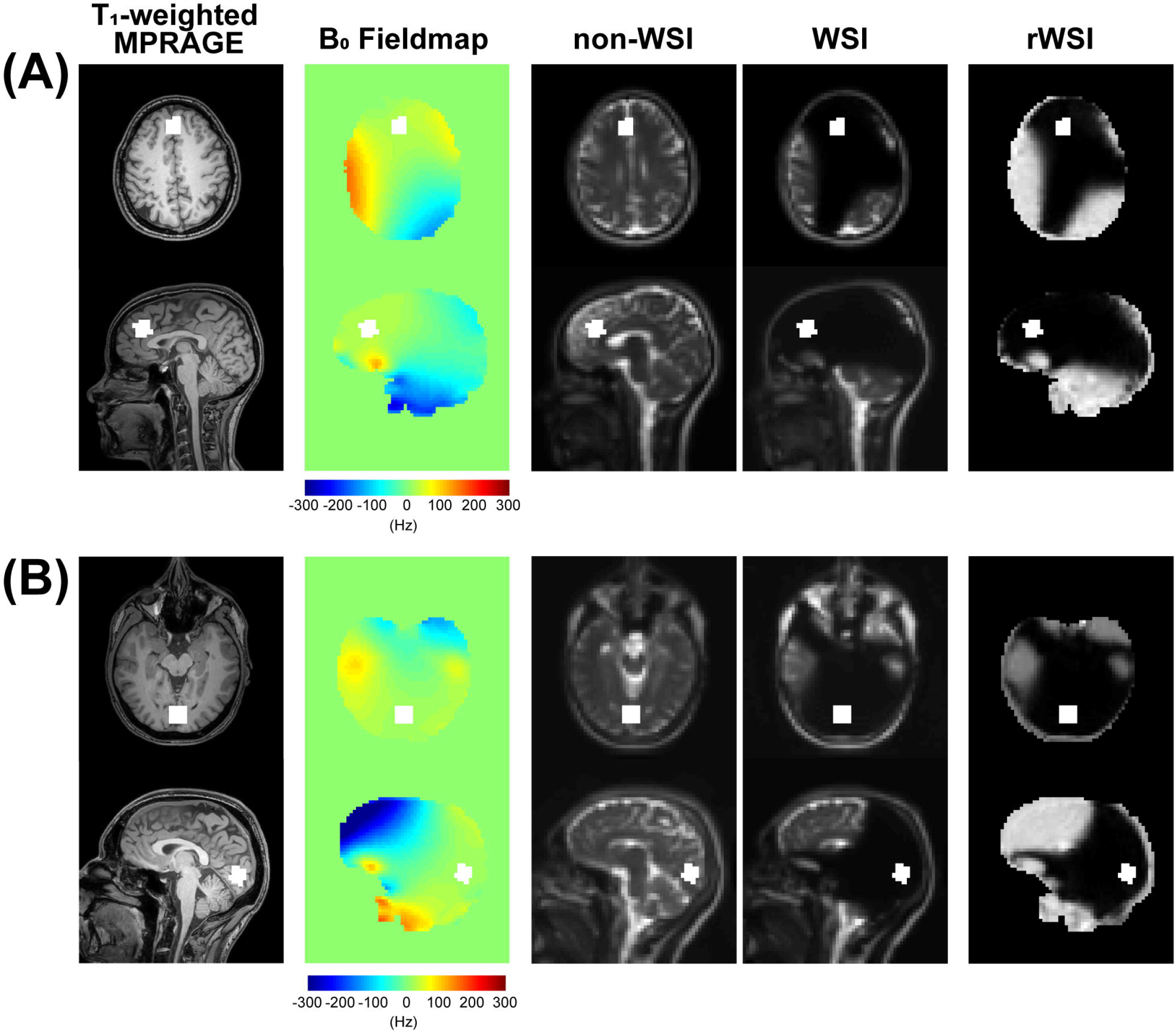
Water-suppression imaging (WSI) for identification of spurious echo artifact regions. Data shown for datasets in ACC (A) and OCC (B).The MRS voxel position is the white box overlaid onto the anatomical T_1_-weighted scan, B_0_-fieldmap, WSI, with and without water suppression (non-WSI). The residual WSI (rWSI) is formed by normalizing the WSI to non-WSI and masking. Note the high similarity between rWSI and B_0_-fieldmap.

### Spectral quality of reconstruction in vivo

Individual channel sensitivity maps, acquired *in vivo* and processed offline, were smoothly varying as expected: representative data is provided in Supporting Information S2. The ERASE reconstruction method in 3 datasets (R03, R06, R02) is shown in Fig. 3. The remaining 3 datasets are provided in Supporting Information S3. Spurious echo artifacts were observed in metabolite spectra in all datasets and regions *(apart from R05)*, and were largely centered over the 3-4 ppm region of the spectra.

Fig. 3A shows artifact estimation and removal results for a voxel placed in the ACC (R03). The rWSI maps revealed unsuppressed water signals in temporoparietal regions of both hemispheres which were automatically segmented into two regions (a, b) and contributed relatively little artifact signal to the spectrum. Two further regions (c and d) adjacent to the frontal sinus were segmented and led to the largest reconstructed artifactual signal components dominated by a signal around 3 ppm. The reconstructed spectrum from the MRS voxel (V) using the ERASE method was visibly improved compared to the standard method, particularly in 3-4ppm region. The appearance of the mI peaks were visibly improved.

Fig. 3B shows results for a voxel in the occipital cortex (R06). The rWSI maps revealed an area of unsuppressed water in the frontal lobe which was automatically segmented into three regions (a,b,c) in addition to a small region near frontal sinus (d) and two more regions inferior to the MRS region in temporal lobe (e) and cerebellum (f). The reconstructed MRS spectrum using the ERASE method was much cleaner than the standard reconstruction. In particular, there was a cleaner appearance of the mI peaks, and complete removal of a spurious peak around 3.3 ppm. The difference between both reconstructions (labeled ‘residual’ in Figure 3) showed no removal of metabolite peaks with ERASE. Lipid contamination was equally present in both the standard and ERASE reconstructions as expected, since no attempt was made to unfold pericranial lipid signals. The temporal lobe region (e) contained relatively little artifactual signal, whereas frontal regions (a) had larger high-frequency artifactual component (>3.5 ppm).

Fig. 3C shows the voxel acquired in the putamen (R02). Standard reconstruction resulted in severe contamination around the choline peak at 3.2 ppm and total creatine peak at 3.9 ppm. Artifactual components were mainly estimated to arise from regions (a and b) both anterior and posterior to the MRS voxel, with large signal over the 3-4 ppm range. ERASE reconstruction revealed a cleaner spectrum over this range, as shown by the improved shape of the choline and creatine peaks.

**Fig. 3:**
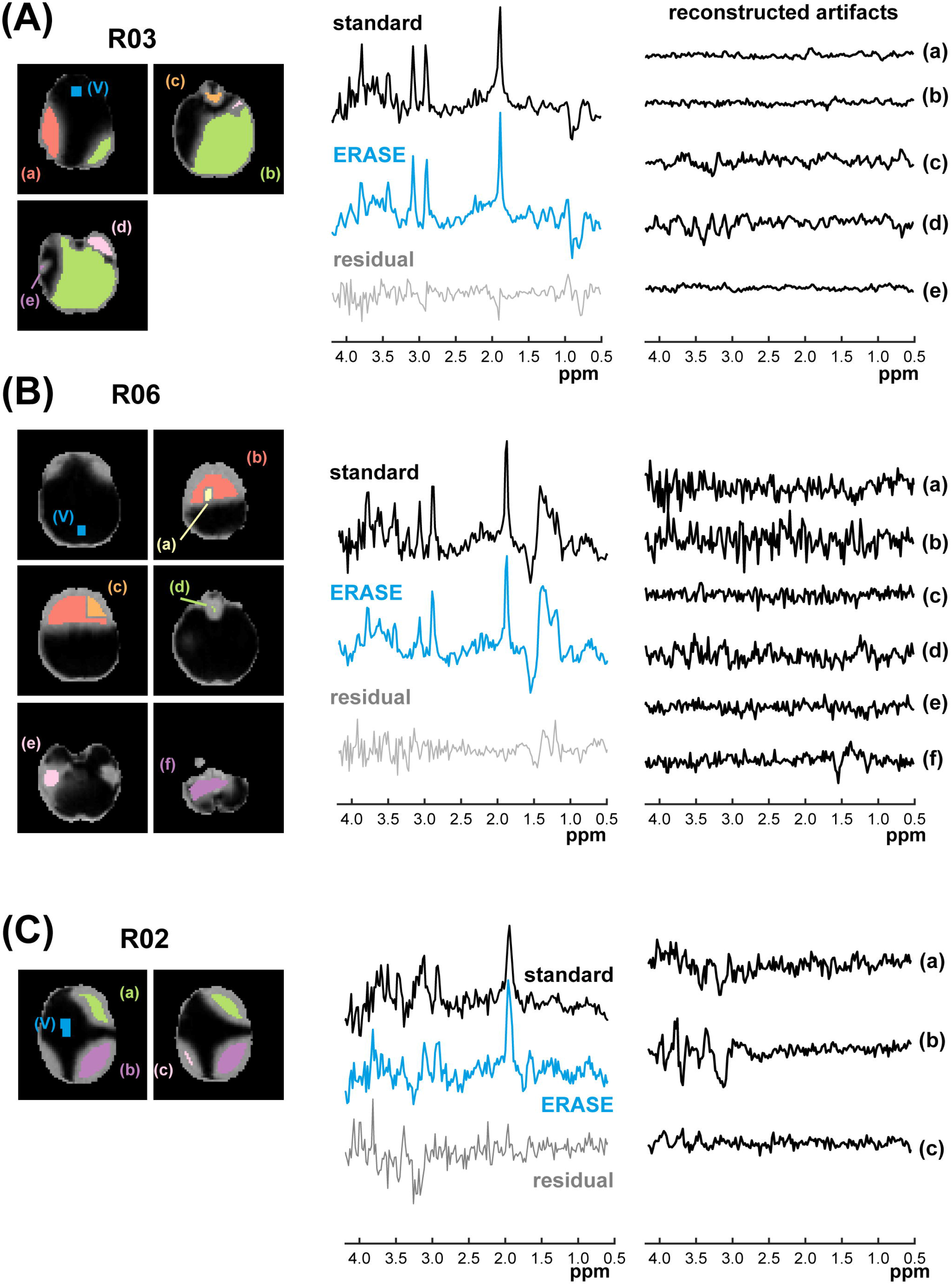
*In vivo* demonstration of the ERASE method in 3 datasets acquired in ACC (A), OCC (B) and putamen (C). Segmented artifact regions are overlaid in color and labelled (a-f) on the rWSI maps for each dataset for different transverse image slices. The MRS voxel (V) is shown in blue. In each dataset the standard reconstruction is shown above the ERASE reconstructed data for the voxel, V (blue). Alongside the metabolite spectra, the artifact spectra reconstructed in each region (a-f) is plotted.

### g-factor and spectral noise

The MRS voxel with the largest associated g-factor (Eq. 3) of reconstruction was dataset R01 (g =2.75) in the frontal region with 8 artifact regions estimated. This dataset also had the largest spectral SNR penalty of reconstruction measured in the spectral domain (54.6 vs. 25.3, standard vs. ERASE), however, still performed well in terms of artifact removal (see Supporting Information S3). The second highest g-factor was for putamen dataset (R02), g = 2.32 with an SNR penalty of (28.6 vs 23.2, standard vs. ERASE). Across remaining datasets, g-factors were lower (1.79, 1.16, 1.14, 1.56). As expected, there was a linear relationship between the g-factor and the SNR penalty of reconstruction (SNR_standard_/SNR_ERASE_) as measured using a regression analysis but this was less than unity (R^2^ = 0.73, β= 0.6, p = 0.03).

### LCModel fitting

Spectra processed with standard combination and ERASE were fit using the LCModel. CRLBs of the five major metabolites are shown in Fig. 4. Across all 6 datasets, CRLBs for tNAA, tCho, tCr, Glx and mI using ERASE were less than or equivalent to standard combination (R05 was unchanged but did not contain significant visible artifacts in the spectrum). Fitting errors were significantly lower using ERASE compared to the standard combination for all major metabolites apart from Glx (11.7% vs. 14.7%, p = 0.06), which was very close to significance; tNAA (5.6% vs. 7.0%, p = 0.03), tCho (10.7% vs.14.1%, p = 0.019), tCr (7.0% vs. 9.2%, p = 0.04) and mI (13.7% vs. 17.5%, p = 0.03). LCModel reported SNR (which accounts for residual unfitted signal in its noise estimation) was significantly improved for ERASE reconstructed spectra than standard combination (8.8 vs. 6.8, p = 0.02, paired t-test).

**Fig. 4:**
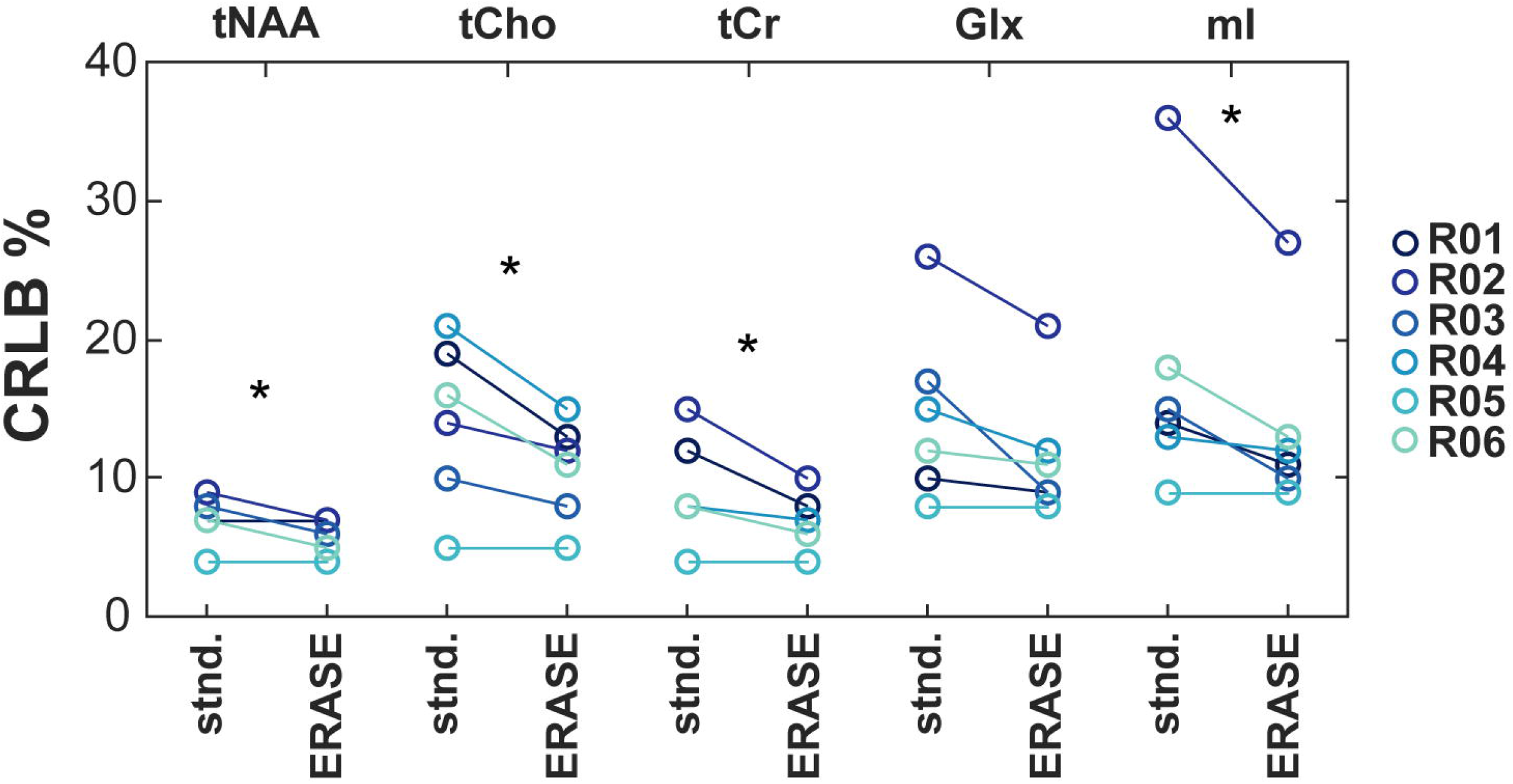
Errors of fitting(CRLBs) for major metabolite signals with standard (stnd.) and ERASE reconstruction for all 6 in vivo datasets after LCModel quantification. (*) indicates p< 0.05 after paired t-test.

## Discussion

An approach for the removal of spurious echo artifacts from single voxel MR spectra is presented, based on receive coil sensitivity profiles. As demonstrated by numerical simulations, the method works best when the spatial origin of spurious echoes is accurately known. In this study, rWSI was used to identify regions likely to contribute artifactual signal; under the conditions used here, frontal regions and the cerebellum were often found to have large amounts of residual water shifted outside the bandwidth of the water suppression pulses. Across 6 *in vivo* datasets acquired in 3 different brain regions, the proposed ERASE method visibly improved spectral quality compared to standard reconstruction of multi-channel MRS data, in particular over the 3-4 ppm region. Spectra reconstructed using ERASE showed lower fitting errors for all major metabolite resonances, reaching statistical significance for tNAA, tCr, tCho, mI (p< 0.05, Fig. 4). Despite the fact that sensitivity-encoding has a g-factor related penalty in SNR, the amount of any SNR reduction was below that attributed to the g-factor alone and did not hinder metabolite fitting in the current examples. It is anticipated that this method will be particularly useful for improving the quality of spectroscopic data when either gradient crushing power is limited, or other factors lead to significant out of voxel magnetization.

In theory, the ERASE method requires only receiver-coil sensitivity maps, which are frequently acquired as a calibration step during a standard MR protocol. Thus, the method could be applied posthoc to data where this information is available. However, unlike previous methods exploiting sensitivity-based unfolding in MRS, such as lipid removal in MRSI (13) and multi-voxel localization (14), the spatial origin of the artifact signal is unknown *a priori*. In order for signal unfolding in Eq. 2 to be valid, sensitivity information must be accurate and the spatial origin of aliased signals known. The numerical phantom simulations (Fig. 1) revealed that when reconstruction region intersects fully with the origin of the artifact, signal separation was complete as in the case of PRIAM (14). Critically for the success of ERASE, partial removal of artifactual signal was achieved even when the reconstruction region did not overlap with artifact origin (Fig. 1D), despite the fact that Eq.2 no longer strictly holds. Partial removal resulted in spectra with improved profile compared to standard reconstruction, provided that the reconstruction regions remained in the same relative position as the underlying origin of the signals.

The WSI technique was relatively straightforward to implement and successfully mapped signal that was not suppressed by the water suppression pulses, revealing potential regions of artifactual signal, similar to data presented in reference (4). Scan time for WSI with whole head coverage was about 2.5 minutes, mainly limited by the TR needed to incorporate the lengthy VAPOR sequence. Together with the non-water suppressed image needed for calculating rWSI maps, the total WSI time was therefore 5 minutes. This could be dramatically reduced in future work, for instance by reducing spatial resolution and using SENSE acceleration. In an MRS study involving multiple spectral locations (and hence multiple high order shim settings), the WSI sequence will need to be repeated for each location. Other approaches could also be considered for identifying regions of unsuppressed water, for instance based on B_0_ maps which showed good concordance with rWSI (e.g. Fig. 2).

Across 6 datasets, independent of voxel geometry, unsuppressed water outside the VOI was often observed in frontal regions. This finding, reflects previous work pointing to frontal sinuses and oral cavity as the main culprit for spurious echo artifacts (8). In addition, due to the use of localized high-order shimming, brain regions remote from the target voxel location often showed large amounts of unsuppressed water, often including the frontal or parietal lobes, and posterior fossa. These regions could potentially be reduced by the use of dynamic B_0_ shimming, with a globally optimized shim set used during water suppression, and a regionally optimized shim set during data acquisition (24,25). WSI could potentially also be used online during scanning to plan acquisition-specific parameters such as order of slice selection gradients or OVS bands,to further reduce these artifacts.

The intensity thresholding and segmentation algorithm applied to rWSI maps generated a small number of volumes for ERASE reconstruction which was limited to a maximum of 8 regions. The theoretical maximum number of regions which can be reconstructed is limited by the total number of receive elements, *K*. However, a large number of volumes will generally decrease the conditioning of the unfolding matrix, hence incur larger g-factor penalties associated with each region. This was observed in dataset R01, where 8 regions were identified (g = 2.75). In this work, low g-factors were measured for reconstruction of the MRS volume, presumably because artifact regions were generally far away from the target MRS volume. A linear relationship was found between g-factors and the ratio of SNR measured using ERASE and standard reconstructions (*β* = 0.6, p = 0.03), yet the influence of g-factor on reconstructed SNR was below the value predicted from SENSE unfolding alone (*β* = 1).This is likely due to two factors: some of the ‘noise’ in the standard reconstruction actually results from very broad residual out-of-voxel signals which are removed by ERASE (e.g. Fig. 3). In addition, the height of the NAA peak used to determine SNR may be altered between reconstructions, and in particular an increase may occur if overlapping signal contributions with opposite phase are removed by ERASE.

A limitation of the ERASE method is that it relies on an assumption that sensitivity of a channel across a reconstruction region is homogeneous, thus sensitivity can be approximated by an average value over the region. This assumption holds for small volumes, since receive profiles are slowly varying. As such, larger volumes for reconstruction will be affected to a greater extent. In this work, the region estimation algorithm was fixed across datasets and did not constrain maximum size, which led to large variation in region volumes (0.09-573 cm^3^). It is suggested that more sophisticated algorithms for region estimation could be implemented, based on a combination of region-growing and optimization by iterating ERASE reconstruction. Constraints could be placed on the total volume size, larger volumes split into sub-volumes, and removal or regions found to contribute no artifact signal.

In conclusion, a new method for reconstruction and removal of spurious echo signals arising from out-of-voxel magnetization is demonstrated. Estimation of the artifact regions was achieved using an imaging-based method and overlapping signal contributions was separated using sensitivity-based reconstruction. This method may improve quantification of metabolites when hardware limitations and voxel geometries lead to contamination of spectra, and could be modified in the future to be run as part of a standard MRS pre-scan.

## Data Availability

Relevant data and functions relating to data processing will be made publicly available on online repository

## Acknowledgments

This work supported in part by NIH P41EB015909. AB would also like to acknowledge the support of the Precision Imaging Beacon, University of Nottingham.

